# Linear regression analysis of COVID-19 outbreak and control in Henan province caused by the output population from Wuhan

**DOI:** 10.1101/2020.05.03.20089193

**Authors:** Cheng Yuan yuan

**Affiliations:** Henan University Huaihe hospital, Kaifeng City, Henan Province, China

**Keywords:** novel coronavirus, COVID-19, Pneumonia, statistical map, epidemic

## Abstract

**Objectives:** To observe outbreak of COVID-19 in Henan province caused by the output population from Wuhan, and high-grade control measures were proformed in Henan province, to study the phase of development and change of the epidemic in Henan province, and to make appropriate inferences about the influence of prevention and control measures and the phase of development of the epidemic.

**Methods:** Linear regression analysis were used to establish a linear regression model with the number of Wuhan roaming people as the dependent variable and the cumulative number of COVID-19 cases in Henan province as the dependent variable, and to calculate and plot the regional distribution of the number of cases in 18 cities in Henan province in accordance with the criteria of whether the number of cases exceeded the expected number.

**Results:** There was a linear correlation between the number of people Wuhan roaming and the number of cases, and the linear regression model equation was statistically significant. The cities that exceeded the expected number of cases had a clear spatio-temporal distribution. Geographically, these cities were roughly in the 1 o’clock and 2 o’clock directions in Nanyang, and in terms of time period, the first phase (10 days), the cities that exceeded the expected number of cases changed almost daily. In the second phase (5 days), cities that exceeded the expected number of cases were moderated, and in the third phase (15 days), cities that exceeded the expected number of cases entered the stabilization phase.

**Conclusions:** The priority cities for COVID-19 prevention and control in Henan province should pay special attention to the cities that have exceeded the expected number of COVID-19 cases, and the implementation of high-level control measures can effectively control the spread of COVID-19 within 2-4 weeks during the early stage of the epidemic.

## Background

From the COVID-19 epidemic area of Wuhan output, the number of people flowing into Henan province, abbreviated as Wuhan roaming. A mobile phone operator has aggregated anonymous data from Wuhan on 22 January 2020 Beijing time (similarly hereinafter), ^[1,2]^ which was the earlier available information on the movement of people from Wuhan into Henan province. There are three base operators in China with a known proportion of subscriber distribution,^[3]^ and assuming a consistent incidence of Wuhan roaming, the results of using Wuhan roaming data from one operator alone, or from all operators, would not affect the linear regression model’s estimate of the expected number of incidences. At the end of 2018, the population of Henan province was 9,605 million, ^[4]^ and Xinyang was the closest city in Henan province to Wuhan, with a distance of about 200 km from Xinyang to Wuhan.

For those departing from Wuhan, it takes only 2-4 hours to enter Henan province. ^[5]^ On January 25, 2020, Henan province announced the initiation of a Level 1 response to a major public health emergency ^[6]^ and began taking the highest level of preventive and control measures. ^[7]^ As of February 23, 2020, a total of 1,271 confirmed cases of COVID-19 were reported in Henan province, of which 19 were deaths. ^[8]^ And from 0000 hours on March 19, 2020, the emergency response level of major public health emergencies in Henan province was lowered from one to two. ^[9]^ We observed the incidence in Henan under Wuhan roaming and high-grade prevention and control measures, using linear regression analysis to classify whether the incidence exceeded the expected number of cases, combined with the cumulative incidence distribution, to study the main phases of development and change of the epidemic in Henan province, and to make appropriate inferences about the impact of prevention and control measures and the phase of development and change of the epidemic.

Statistical analysis software: R software, version 3.6.3.

## Methods

The linear regression model is *Y=α+βX*, whose estimation model is *Ŷ = a + bX*, with Wuhan roaming as the dependent variable *X*, and the cumulative daily number of cases in 18 cities in Henan province from DAY 1 to DAY 30 as the dependent variable *Y*. DAY 1 to DAY 30 corresponds to the cumulative number of cases in Henan province from January 25 to February 23, 2020. ^[11–40]^

Take *X* as the horizontal coordinate, and *Y* as the vertical coordinate to draw a scatterplot.

Statistical mapping is carried out using the criteria of whether the actual number of cases exceeded the expected number of cases.

The days is used as the horizontal coordinate, and the number of cumulative cases in the city is used as the vertical coordinate to draw a line graph of cumulative cases.

## Results

### Scatter plots and linear regression models

Linear regression analysis was performed using DAY 1 as an example, and the linear analysis and plotting from DAY 2 to DAY 30 in the same way as DAY 1.

Using *X* as the horizontal coordinate and *Y* = DAY 1 as the vertical coordinate, a scatterplot was drawn, see Figure 1(a). The outliers were removed (Zhengzhou cumulative number of cases), and a scatterplot was made again, see Figure 1(b), to build a linear regression model with the data after removed the outliers.

**Figures 1:**
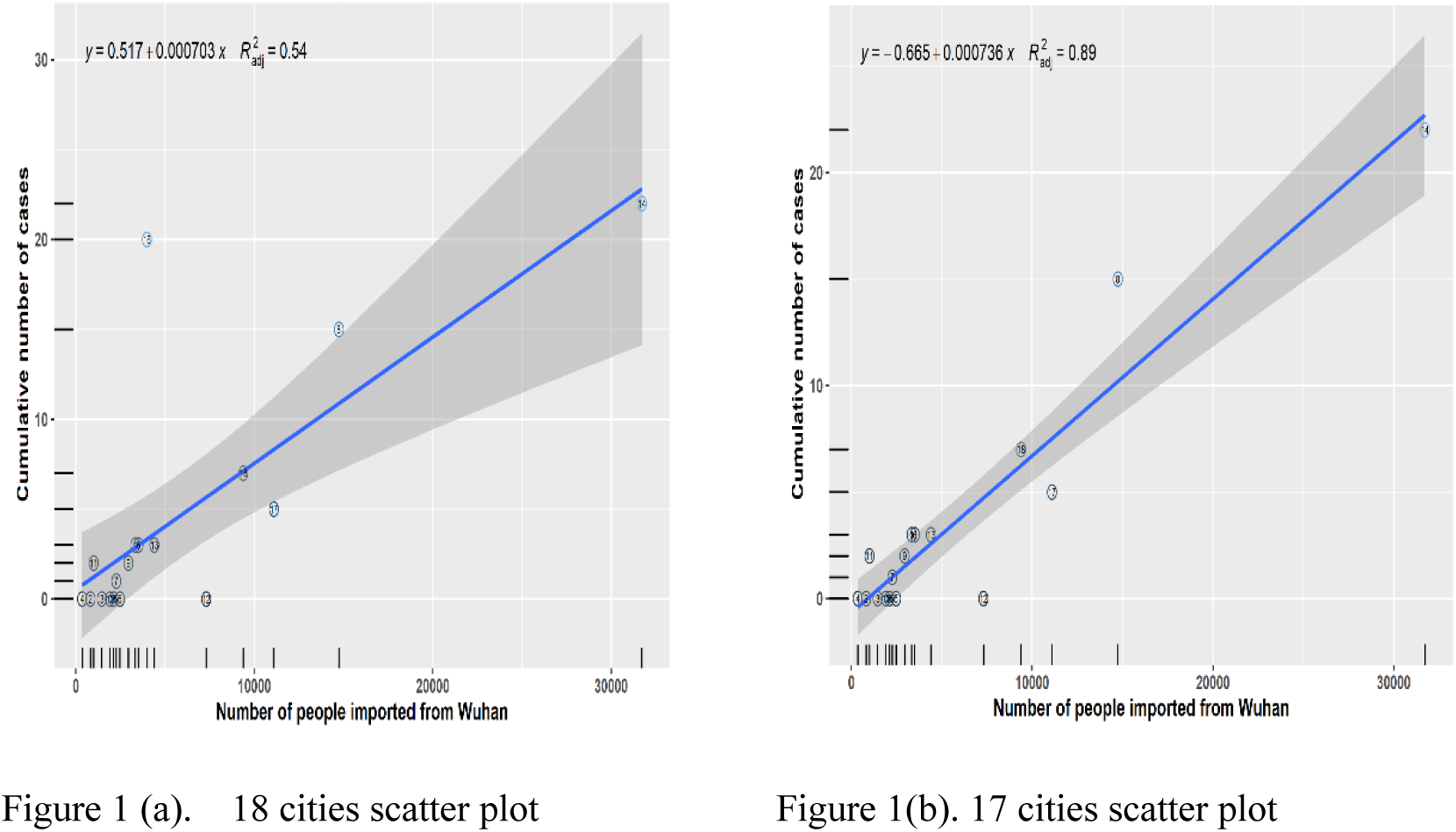
Scatter plots of Wuhan roaming and cumulative number of cases in Henan. Figures 1(a) was 18 cities scatter plot, Figures 1(b) was 17 cities scatter plot which removing outliers. Figures 1. showed that there were a linear correlation between Wuhan roaming and cumulative number of cases in Henan.

Residual plots analysis were shown in Figures 2(a) and Figure 2(b).

**Figures 2:**
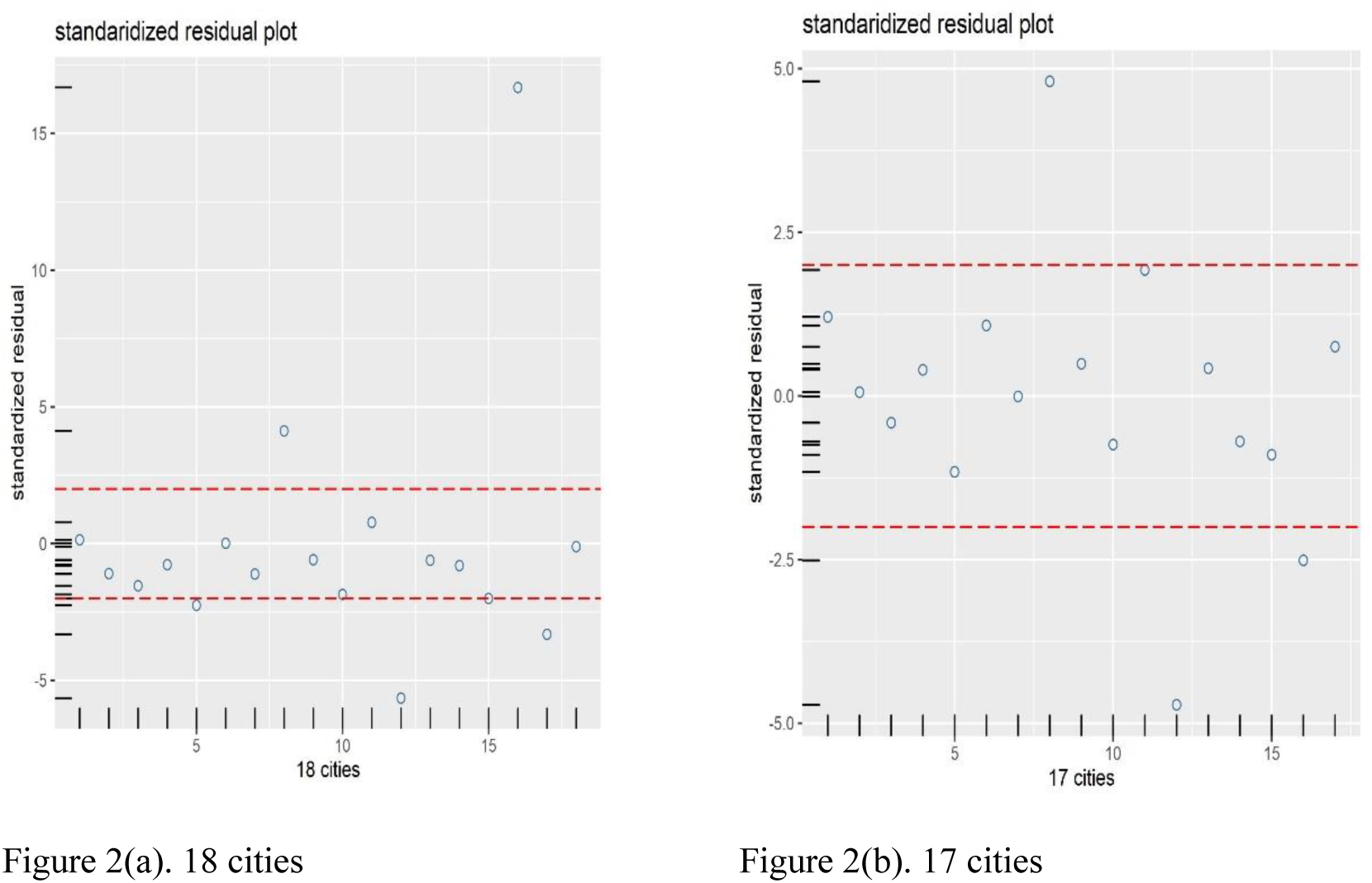
Standaridized residual plots of linear regression models. Figure 2(a) was standaridized residual plots of 18 cities in Henan, and Figure 2(b) was standaridized residual plots of 17 cities in Henan. Figure 2(a) and Figure 2(b) show that the scatter was evenly distributed above and below the zero reference line, mostly between ±2 times the standard deviation, and the model fited well.

The correlation coefficient of the linear regression model = 0.945, decision coefficient = 0.893, adjusted decision coefficient = 0.886, F-value = 125.6, P-value = 1.096e-08. The model equation was statistically significant. The regression equation model was: *Ŷ = -0.6645+0.0007X*, the value of *X* was substituted into the above equation, the expected number of cases was calculated, and the expected number of cases was used to calculate the ratio of expected cases, exceeded the expected number of cases. The calculation results were shown in Table 1.

**Table 1.**
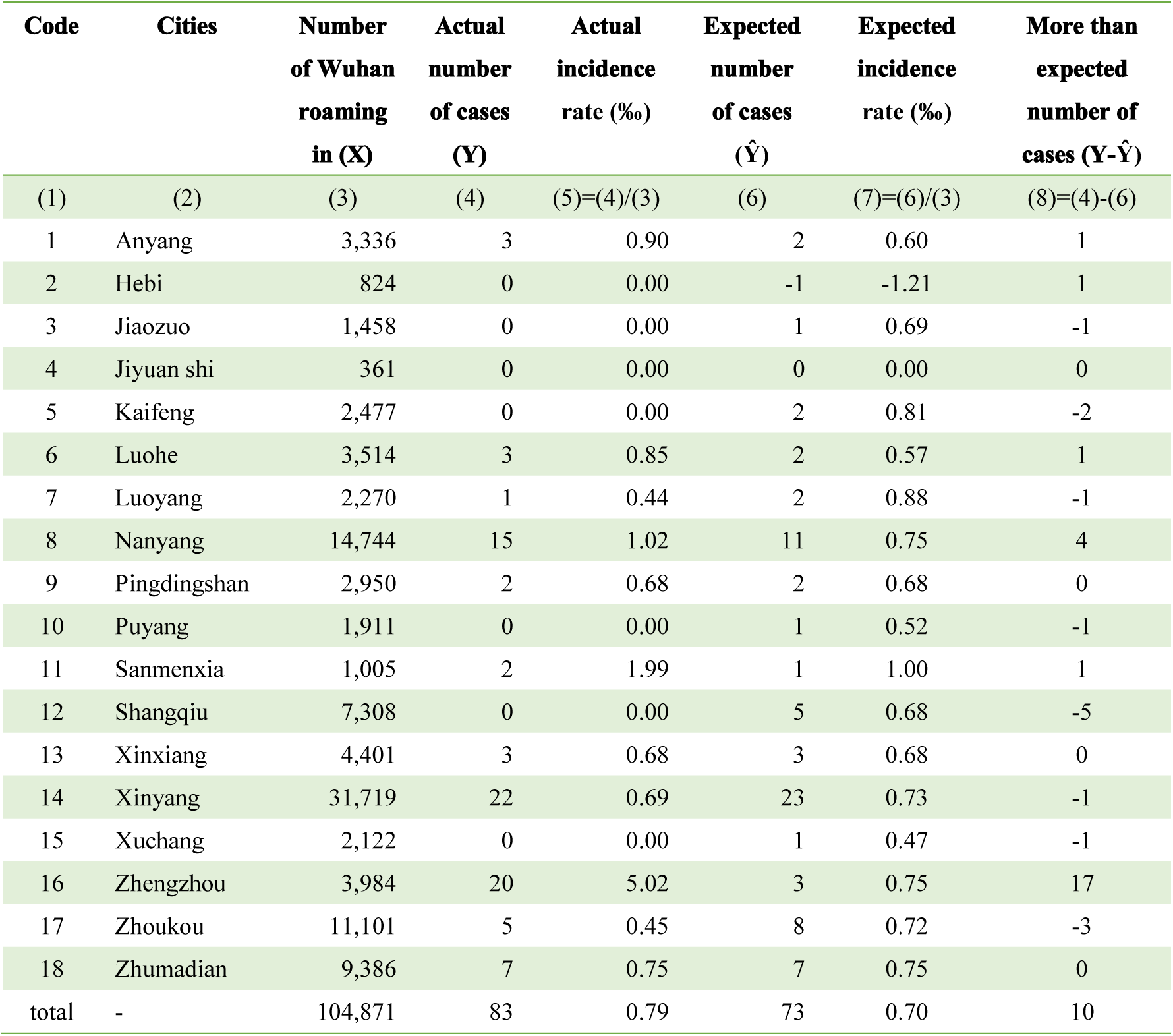
Data on the number of COVID-19 cases in Henan in Day 1.

Nine cities (city code: 16, 8, 11, 1, 6, 18, 9, 13, and 2) exceed the expected number of cases, while the remaining nine cities did not exceed the expected number of cases. The statistical map was drawn according to the criteria of whether the cumulative number of cases in 18 cities in Henan province exceeded the expected number of cases on the same day. DAY 2 - DAY 30 was calculated and plotted in the same way as DAY 1. Data on the number of COVID-19 cases in Henan in 30 days (Table 2), and data on the exceeded expected number of COVID-19 cases in Henan in 30 days (Table 3). Statistical map see Figure 3.

**Table 2.** Data on the number of COVID-19 cases in Henan in 30 days.

| Code | Actual number of cases (Y) |  |  |  |  |  |  |  |  |  |  |  |  |  |  |  |  |  |  |  |  |  |  |  |  |  |  |  |  |  |
| --- | --- | --- | --- | --- | --- | --- | --- | --- | --- | --- | --- | --- | --- | --- | --- | --- | --- | --- | --- | --- | --- | --- | --- | --- | --- | --- | --- | --- | --- | --- |
|  | DAY1 | DAY2 | DAY3 | DAY4 | DAY5 | DAY6 | DAY7 | DAY8 | DAY9 | DAY10 | DAY11 | DAY12 | DAY13 | DAY14 | DAY15 | DAY16 | DAY17 | DAY18 | DAY19 | DAY20 | DAY21 | DAY22 | DAY23 | DAY24 | DAY25 | DAY26 | DAY27 | DAY28 | DAY29 | DAY30 |
| 1 | 3 | 7 | 11 | 14 | 17 | 21 | 25 | 25 | 29 | 33 | 35 | 37 | 40 | 43 | 44 | 45 | 46 | 49 | 50 | 50 | 51 | 51 | 51 | 52 | 52 | 53 | 53 | 53 | 53 | 53 |
| 2 | 0 | 2 | 2 | 5 | 6 | 6 | 6 | 8 | 8 | 10 | 10 | 10 | 11 | 13 | 17 | 17 | 18 | 19 | 19 | 19 | 19 | 19 | 19 | 19 | 19 | 19 | 19 | 19 | 19 | 19 |
| 3 | 0 | 0 | 2 | 2 | 2 | 2 | 2 | 7 | 8 | 12 | 15 | 17 | 18 | 20 | 24 | 25 | 28 | 28 | 29 | 29 | 30 | 30 | 32 | 32 | 32 | 32 | 32 | 32 | 32 | 32 |
| 4 | 0 | 0 | 0 | 0 | 0 | 0 | 0 | 0 | 2 | 2 | 2 | 3 | 4 | 4 | 4 | 4 | 4 | 4 | 4 | 4 | 4 | 4 | 5 | 5 | 5 | 5 | 5 | 5 | 5 | 5 |
| 5 | 0 | 1 | 2 | 3 | 4 | 6 | 8 | 11 | 11 | 12 | 17 | 18 | 20 | 21 | 24 | 24 | 25 | 25 | 25 | 25 | 25 | 26 | 26 | 26 | 26 | 26 | 26 | 26 | 26 | 26 |
| 6 | 3 | 6 | 9 | 9 | 11 | 13 | 14 | 16 | 20 | 23 | 24 | 26 | 26 | 29 | 30 | 30 | 31 | 33 | 33 | 33 | 34 | 34 | 34 | 34 | 34 | 35 | 35 | 35 | 35 | 35 |
| 7 | 1 | 2 | 2 | 2 | 2 | 3 | 5 | 7 | 7 | 12 | 17 | 21 | 24 | 25 | 25 | 27 | 28 | 29 | 30 | 30 | 31 | 31 | 31 | 31 | 31 | 31 | 31 | 31 | 31 | 31 |
| 8 | 15 | 19 | 26 | 31 | 51 | 61 | 66 | 76 | 84 | 99 | 107 | 111 | 118 | 128 | 133 | 134 | 136 | 138 | 145 | 146 | 150 | 151 | 154 | 155 | 155 | 155 | 155 | 155 | 155 | 155 |
| 9 | 2 | 5 | 5 | 5 | 8 | 10 | 11 | 14 | 19 | 28 | 34 | 41 | 46 | 48 | 50 | 52 | 54 | 55 | 56 | 57 | 58 | 58 | 58 | 58 | 58 | 58 | 58 | 58 | 58 | 58 |
| 10 | 0 | 0 | 0 | 1 | 1 | 3 | 3 | 3 | 3 | 4 | 4 | 5 | 5 | 7 | 9 | 10 | 11 | 13 | 13 | 13 | 17 | 17 | 17 | 17 | 17 | 17 | 17 | 17 | 17 | 17 |
| 11 | 2 | 3 | 3 | 4 | 5 | 6 | 7 | 7 | 7 | 7 | 7 | 7 | 7 | 7 | 7 | 7 | 7 | 7 | 7 | 7 | 7 | 7 | 7 | 7 | 7 | 7 | 7 | 7 | 7 | 7 |
| 12 | 0 | 6 | 7 | 14 | 18 | 31 | 36 | 44 | 50 | 60 | 61 | 65 | 72 | 76 | 82 | 83 | 86 | 86 | 87 | 88 | 90 | 91 | 91 | 91 | 91 | 91 | 91 | 91 | 91 | 91 |
| 13 | 3 | 4 | 6 | 7 | 12 | 17 | 23 | 23 | 27 | 33 | 39 | 41 | 41 | 43 | 44 | 46 | 48 | 53 | 54 | 55 | 55 | 56 | 57 | 57 | 57 | 57 | 57 | 57 | 57 | 57 |
| 14 | 22 | 23 | 29 | 32 | 42 | 49 | 70 | 88 | 99 | 112 | 138 | 164 | 176 | 192 | 205 | 220 | 228 | 231 | 240 | 243 | 252 | 261 | 262 | 266 | 268 | 269 | 270 | 273 | 274 | 274 |
| 15 | 0 | 1 | 2 | 2 | 3 | 8 | 12 | 13 | 15 | 19 | 24 | 26 | 28 | 30 | 31 | 31 | 32 | 34 | 34 | 37 | 37 | 38 | 38 | 38 | 39 | 39 | 39 | 39 | 39 | 39 |
| 16 | 20 | 29 | 37 | 40 | 46 | 50 | 56 | 65 | 72 | 85 | 92 | 102 | 112 | 120 | 126 | 130 | 132 | 137 | 141 | 142 | 144 | 148 | 151 | 154 | 156 | 156 | 157 | 157 | 157 | 157 |
| 17 | 5 | 11 | 15 | 19 | 25 | 36 | 38 | 40 | 47 | 52 | 56 | 59 | 60 | 62 | 62 | 65 | 65 | 66 | 68 | 69 | 70 | 71 | 75 | 76 | 76 | 76 | 76 | 76 | 76 | 76 |
| 18 | 7 | 9 | 10 | 16 | 25 | 30 | 40 | 46 | 58 | 72 | 82 | 98 | 106 | 113 | 116 | 123 | 126 | 128 | 134 | 137 | 138 | 138 | 138 | 139 | 139 | 139 | 139 | 139 | 139 | 139 |

**Table 3.** Data on the exceeded expected number of COVID-19 cases in Henan in 30 days.

| Code | Expected number of cases ($\hat{Y}$) | | | | | | | | | | | | | | | | | | | | | | | | | | | | | |
| --- | --- | --- | --- | --- | --- | --- | --- | --- | --- | --- | --- | --- | --- | --- | --- | --- | --- | --- | --- | --- | --- | --- | --- | --- | --- | --- | --- | --- | --- | --- |
|  | DAY1 | DAY2 | DAY3 | DAY4 | DAY5 | DAY6 | DAY7 | DAY8 | DAY9 | DAY10 | DAY11 | DAY12 | DAY13 | DAY14 | DAY15 | DAY16 | DAY17 | DAY18 | DAY19 | DAY20 | DAY21 | DAY22 | DAY23 | DAY24 | DAY25 | DAY26 | DAY27 | DAY28 | DAY29 | DAY30 |
| 1 | 1.2 | 3.2 | 5.9 | 7.2 | 7.6 | 8.2 | 10.0 | 7.8 | 8.9 | 8.4 | 7.5 | 6.8 | 7.7 | 8.5 | 7.6 | 7.6 | 7.4 | 9.1 | 9.1 | 8.5 | 8.7 | 8.4 | 7.9 | 8.7 | 8.7 | 9.6 | 9.6 | 9.6 | 9.6 | 9.6 |
| 2 | 0.1 | 0.2 | -0.6 | 1.1 | 0.6 | -2.0 | -2.7 | -1.5 | -3.4 | -4.8 | -5.9 | -6.7 | -7.0 | -5.9 | -2.9 | -2.9 | -2.6 | -2.8 | -2.9 | -3.3 | -3.5 | -3.1 | -3.5 | -3.4 | -3.3 | -3.4 | -3.3 | -3.0 | -3.0 | -3.0 |
| 3 | -0.4 | -2.3 | -1.2 | -2.6 | -4.4 | -7.2 | -8.3 | -4.5 | -5.6 | -5.3 | -3.9 | -3.1 | -3.6 | -2.9 | -0.1 | 0.7 | 2.8 | 1.6 | 2.3 | 1.9 | 2.5 | 2.7 | 4.3 | 4.4 | 4.4 | 4.3 | 4.4 | 4.6 | 4.6 | 4.6 |
| 4 | 0.0 | -1.4 | -2.1 | -3.4 | -4.6 | -7.1 | -7.5 | -8.1 | -7.8 | -11.0 | -11.8 | -11.3 | -11.3 | -12.1 | -12.9 | -12.7 | -13.3 | -14.4 | -14.4 | -14.7 | -14.8 | -14.3 | -13.7 | -13.5 | -13.4 | -13.5 | -13.4 | -13.1 | -13.0 | -13.0 |
| 5 | -1.2 | -2.1 | -2.2 | -2.8 | -4.1 | -5.1 | -4.8 | -3.6 | -6.1 | -9.2 | -6.6 | -7.6 | -7.4 | -8.2 | -6.7 | -7.4 | -7.5 | -8.7 | -9.4 | -9.9 | -10.5 | -9.6 | -10.1 | -10.1 | -10.1 | -10.2 | -10.2 | -10.1 | -10.1 | -10.1 |
| 6 | 1.1 | 2.1 | 3.7 | 2.0 | 1.3 | -0.1 | -1.4 | -1.8 | -0.7 | -2.3 | -4.4 | -5.1 | -7.4 | -6.6 | -7.5 | -8.6 | -8.9 | -8.2 | -9.2 | -9.8 | -9.7 | -10.0 | -10.6 | -10.7 | -10.8 | -9.9 | -9.9 | -9.9 | -9.9 | -9.9 |
| 7 | 0.0 | -0.9 | -2.0 | -3.6 | -5.7 | -7.7 | -7.3 | -7.0 | -9.4 | -8.4 | -5.6 | -3.5 | -2.2 | -2.9 | -4.4 | -3.0 | -3.0 | -3.2 | -2.8 | -3.3 | -2.9 | -2.9 | -3.4 | -3.4 | -3.4 | -3.5 | -3.4 | -3.3 | -3.3 | -3.3 |
| 8 | 4.8 | 6.2 | 9.5 | 11.2 | 23.1 | 26.4 | 22.3 | 23.9 | 24.5 | 30.0 | 26.8 | 19.8 | 20.5 | 22.7 | 22.0 | 17.2 | 15.7 | 15.7 | 18.1 | 17.4 | 17.7 | 15.6 | 17.4 | 16.8 | 16.2 | 15.8 | 15.5 | 14.7 | 14.4 | 14.4 |
| 9 | 0.5 | 1.5 | 0.3 | -1.4 | -0.8 | -2.0 | -3.0 | -2.0 | 0.2 | 4.9 | 8.3 | 12.9 | 15.9 | 15.9 | 16.2 | 17.3 | 18.1 | 17.9 | 18.0 | 18.5 | 18.7 | 18.6 | 18.1 | 17.9 | 17.9 | 17.8 | 17.8 | 17.9 | 17.9 | 17.9 |
| 10 | -0.7 | -2.6 | -3.7 | -4.2 | -6.1 | -7.1 | -8.4 | -9.9 | -12.2 | -15.0 | -16.9 | -17.5 | -19.2 | -18.7 | -18.0 | -17.5 | -17.4 | -16.6 | -17.1 | -17.6 | -14.1 | -14.0 | -14.4 | -14.4 | -14.4 | -14.5 | -14.4 | -14.3 | -14.2 | -14.2 |
| 11 | 1.9 | 1.1 | 0.2 | -0.1 | -0.7 | -2.3 | -2.1 | -3.1 | -5.1 | -8.5 | -9.8 | -10.7 | -12.0 | -13.1 | -14.1 | -14.1 | -14.9 | -16.1 | -16.3 | -16.7 | -16.9 | -16.6 | -17.0 | -16.9 | -16.8 | -16.9 | -16.8 | -16.6 | -16.5 | -16.5 |
| 12 | -4.7 | -0.9 | -2.1 | 2.7 | 2.1 | 10.6 | 11.0 | 14.6 | 16.2 | 19.9 | 15.1 | 13.6 | 17.0 | 16.8 | 19.7 | 18.0 | 18.9 | 17.4 | 16.2 | 16.2 | 16.3 | 16.1 | 15.3 | 14.7 | 14.4 | 14.2 | 14.1 | 13.9 | 13.8 | 13.8 |
| 13 | 0.4 | -0.6 | -0.2 | -1.0 | 0.8 | 2.2 | 5.3 | 2.5 | 3.2 | 4.3 | 6.6 | 5.1 | 2.6 | 1.9 | 0.7 | 1.2 | 1.7 | 5.4 | 5.1 | 5.4 | 4.3 | 4.8 | 5.2 | 4.9 | 4.8 | 4.6 | 4.6 | 4.6 | 4.5 | 4.5 |
| 14 | -0.7 | -3.2 | -4.6 | -7.3 | -13.3 | -18.2 | -16.4 | -16.1 | -19.0 | -23.2 | -20.6 | -18.0 | -18.4 | -18.6 | -17.0 | -15.0 | -13.9 | -14.0 | -15.0 | -15.2 | -14.3 | -12.6 | -13.8 | -13.4 | -12.9 | -12.6 | -12.3 | -11.6 | -11.3 | -11.3 |
| 15 | -0.9 | -1.8 | -1.9 | -3.4 | -4.5 | -2.5 | 0.1 | -0.5 | -0.9 | -0.8 | 2.1 | 2.3 | 2.6 | 3.0 | 2.6 | 2.1 | 2.1 | 2.8 | 2.3 | 4.8 | 4.3 | 5.3 | 4.8 | 4.8 | 5.9 | 5.8 | 5.8 | 5.9 | 6.0 | 6.0 |
| 16 | 17.7 | 24.7 | 31.3 | 32.5 | 35.5 | 36.0 | 39.4 | 45.8 | 49.7 | 57.9 | 61.5 | 68.4 | 76.0 | 81.5 | 85.4 | 88.1 | 88.7 | 92.4 | 95.2 | 95.6 | 96.6 | 100.2 | 102.6 | 105.3 | 107.3 | 107.1 | 108.1 | 108.1 | 108.1 | 108.1 |
| 17 | -2.5 | 1.1 | 2.1 | 3.3 | 3.0 | 8.3 | 3.5 | -1.0 | 0.1 | -2.8 | -7.4 | -12.7 | -16.7 | -20.7 | -25.1 | -26.4 | -29.2 | -30.0 | -31.5 | -31.7 | -33.6 | -34.8 | -31.8 | -31.9 | -32.3 | -32.6 | -32.8 | -33.4 | -33.6 | -33.6 |
| 18 | 0.8 | 0.4 | -1.2 | 2.3 | 5.8 | 5.6 | 9.8 | 10.3 | 17.0 | 23.8 | 26.5 | 35.5 | 39.1 | 40.9 | 40.1 | 43.5 | 44.0 | 44.4 | 47.5 | 49.4 | 47.9 | 46.2 | 45.3 | 45.4 | 45.1 | 44.8 | 44.7 | 44.2 | 44.0 | 44.0 |

The cities in Henan province that exceeded the expected number of cases were:

Figure 3(a). DAY 1: Zhengzhou, Nanyang, Sanmenxia, Anyang, Luohe, Zhumadian, Pingdingshan, Xinxiang, and Hebi (cities code: 16, 8, 11, 1, 6, 18, 9, 13, and 2);

Figure 3(b). DAY 2: Zhengzhou, Nanyang, Anyang, Luohe, Pingdingshan, Zhoukou, Sanmenxia, Zhumadian, and Hebi (cities code: 16, 8, 1, 6, 9, 17, 11, 18, and 2);

Figure 3(c). DAY 3: Zhengzhou, Nanyang, Anyang, Luohe, Zhoukou, Pingdingshan, and Sanmenxia (cities code: 16, 8, 1, 6, 17, 9, and 11);

Figure 3 (d). DAY 4: Zhengzhou, Nanyang, Anyang, Zhoukou, Shangqiu, Zhumadian, Luohe, and Hebi (cities code: 16, 8, 1, 17, 12, 18, 6, and 2);

Figure 3(e). DAY 5: Zhengzhou, Nanyang, Anyang, Zhumadian, Zhoukou, Shangqiu, Luohe, Xinxiang, and Hebi (cities code: 16, 8, 1, 18, 17, 12, 6, 13, and 2);

Figure 3(f). DAY 6: Zhengzhou, Nanyang, Shangqiu, Zhoukou, Anyang, Zhumadian, and Xinxiang (cities code: 16, 8, 12, 17, 1, 18, and 13);

Figure 3(g). DAY 7: Zhengzhou, Nanyang, Shangqiu, Anyang, Zhumadian, Xinxiang, Zhoukou, and Xuchang (cities code: 16, 8, 12, 1, 18, 13, 17, and 15);

Figure 3(h). DAY 8: Zhengzhou, Nanyang, Shangqiu, Zhumadian, Anyang, and Xinxiang (cities code: 16, 8, 12, 18, 1, and 13);

Figure 3(i). DAY 9: Zhengzhou, Nanyang, Zhumadian, Shangqiu, Anyang, Xinxiang, Pingdingshan, and Zhoukou (cities code: 16, 8, 18, 12, 1, 13, 9, and 17);

Figure 3(j). DAY 10: Zhengzhou, Nanyang, Zhumadian, Shangqiu, Anyang, Pingdingshan, and Xinxiang (cities code: 16, 8, 18, 12, 1, 9, and 13);

Figure 3(k). DAY 11-15: Zhengzhou, Nanyang, Zhumadian, Shangqiu, Pingdingshan, Anyang, Xinxiang, and Xuchang (cities code: 16, 8, 18, 12, 9, 1, 13, and 15);

Figure 3 (l). DAY 16-30: Zhengzhou, Zhumadian, Shangqiu, Pingdingshan, Nanyang, Anyang, Xuchang, Xinxiang, and Jiaozuo (cities code: 16, 18, 12, 9, 8, 1, 15, 13, and 3). With the remaining cities not exceeded the expected number of cases.

**Figure 3:**
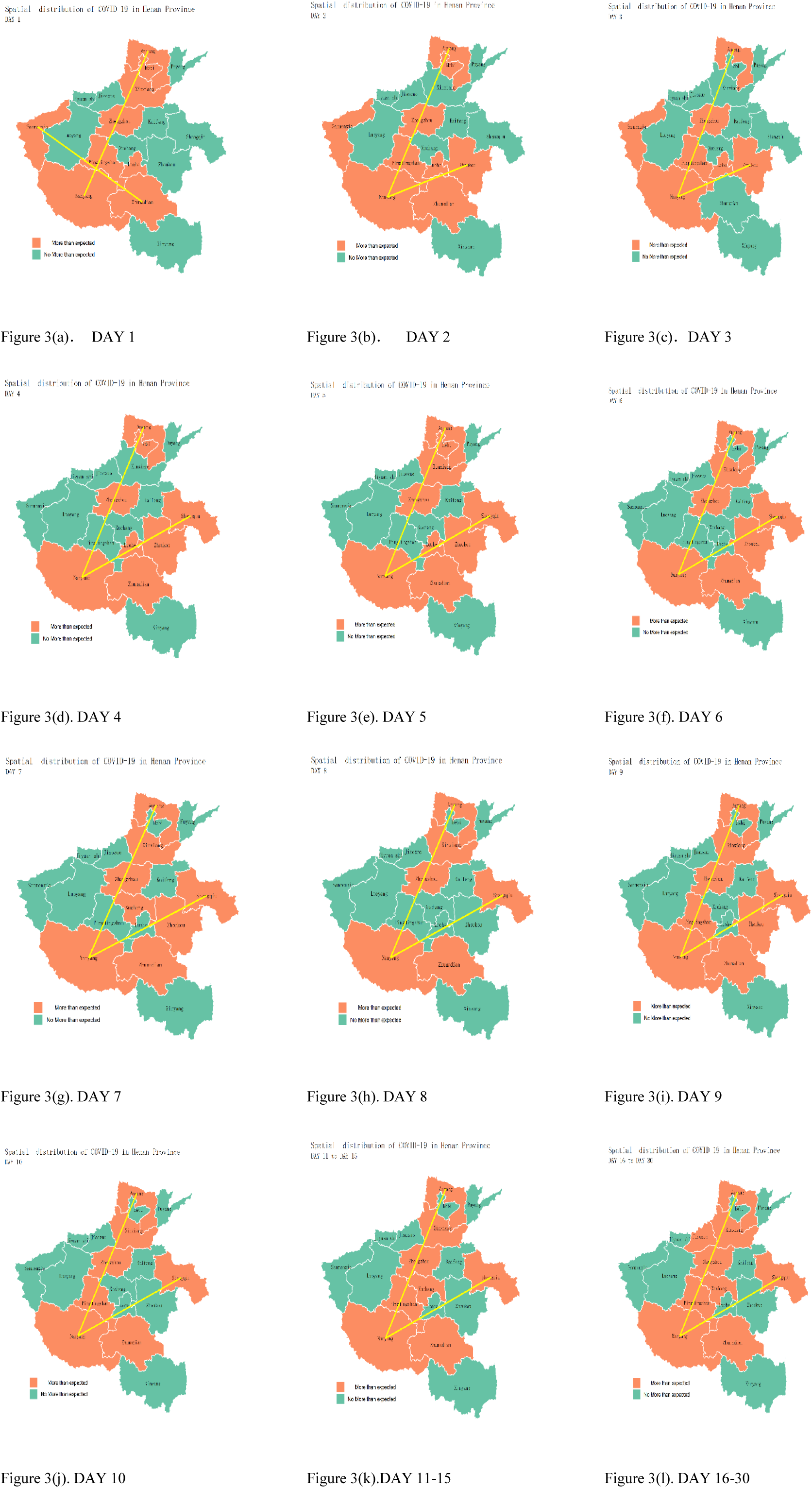
Statistical map of cities with exceeded the expected cumulative number of cases in 30 days.

There were three main phases of disease development in terms of time distribution, with DAY 1 - DAY10 as the first phase, DAY 11 - DAY 15 as the second phase, and DAY 16 - DAY 30 as the third phase.

The geographical distribution of the disease was mainly as follows: cities with more than the expected number of cases were mainly distributed in the 1 o’clock and 2 o’clock directions with Nanyang as the midpoint. The 1 o’clock direction covered six cities, including Nanyang, Pingdingshan, Zhengzhou, Xinxiang, Hebi, and Anyang. The 2 o’clock direction covers four cities, including Zhumadian, Luohe, Zhoukou, and Shangqiu.

### Line plot

Cumulative number of cases for 30 days see Figure 4(a). Expected number of cases for 30 days see Figure 4(b).

**Figure 4:**
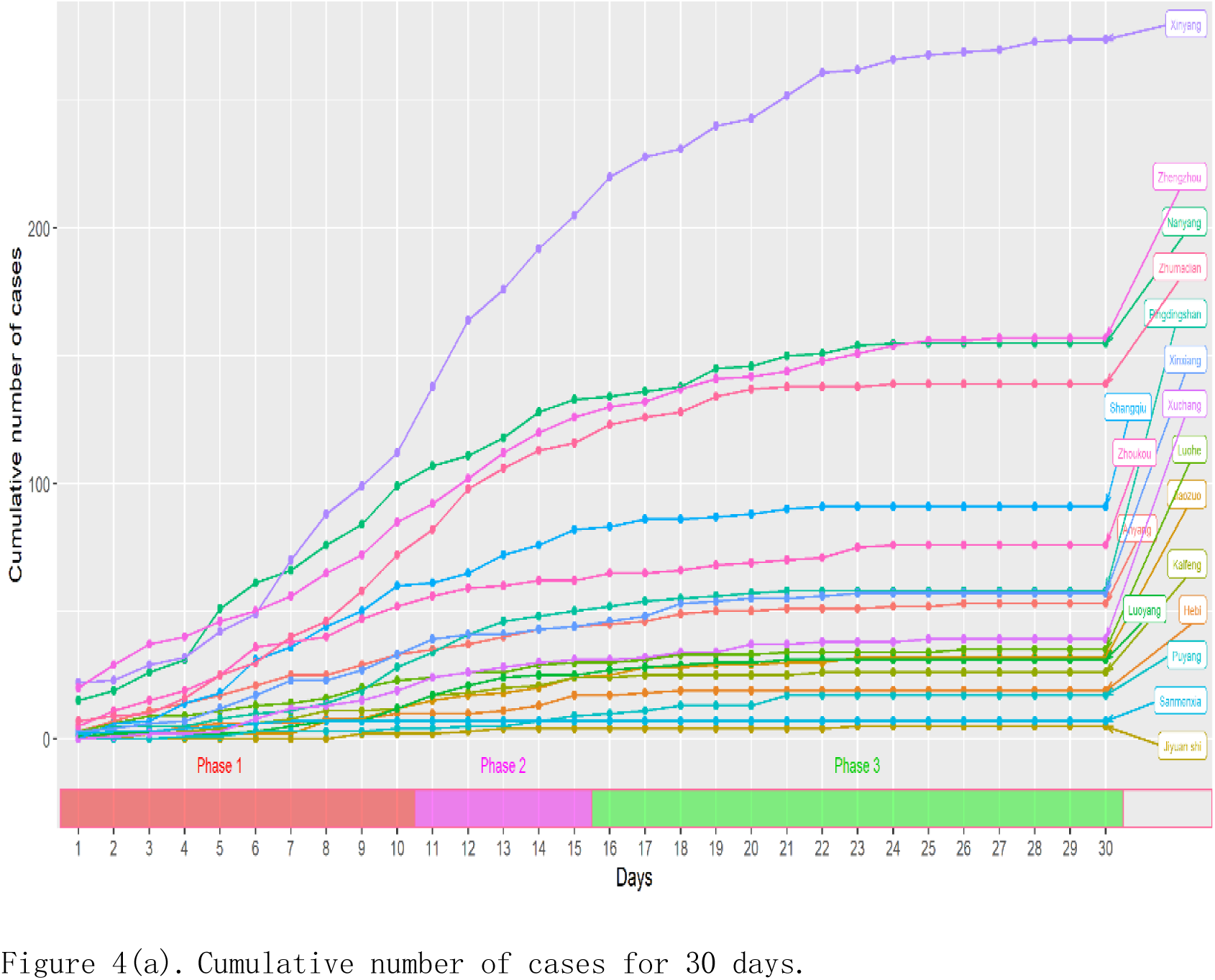
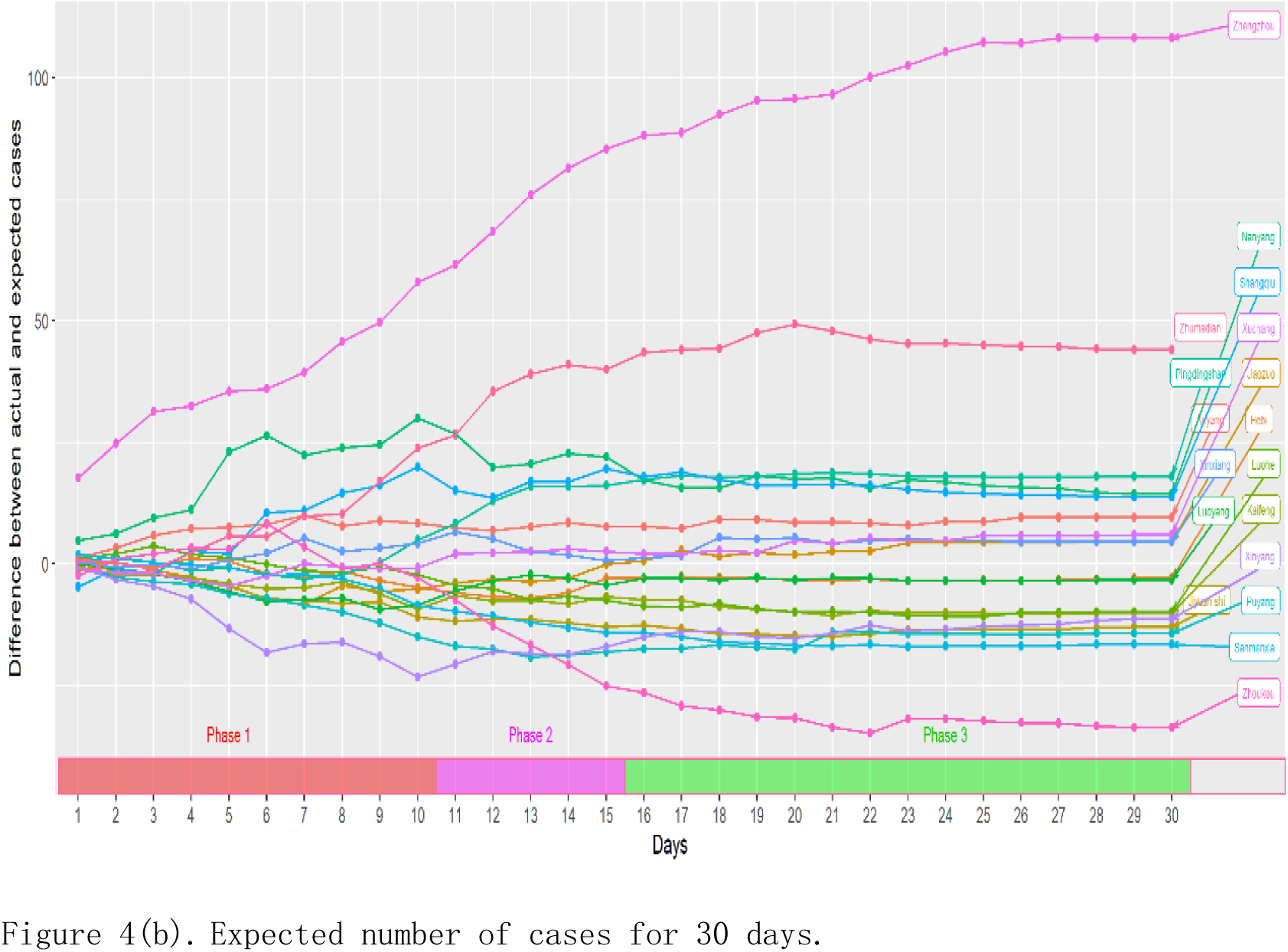
Line plot of cumulative and expected number of cases. Figure 4(a) showed that after three phases of development, the elevation angle of the cumulative onset curve gradually shifted from upward to downward pressure and tended to level out. Figure 4(b) showed that after three phases of development, most urban morbidity curves have shifted from dramatic fluctuations to smooth and from clutter to order.

As can be seen from Figure 3 and Figure 4, the list of cities that exceeded the expected number of cases in the first phase (DAY 1 - DAY 10, 10 days) changed daily, and the incidence curve was highly volatile and chaotic, with a faster increase in the cumulative incidence curve and more dramatic changes in the outbreak. In the second phase (DAY 11 - DAY 15, 5 days), the cities that exceeded the expected number of cases were fixed in 8 cities, including Zhengzhou, Nanyang, Zhumadian, Shangqiu, Pingdingshan, Anyang, Xinxiang, and Xuchang (cities code: 16, 8, 18, 12, 9, 1, 13, and 15). The incidence curve showed that it entered the combing phase, the elevation angle of the cumulative incidence curve was downwardly pressed, and the level of incidence was generally suppressed, showing a certain relief. After 10 days of strict intervention, the epidemic was effectively suppressed, after which it entered the combing phase five days later and, after minor fluctuations, the control phase.

## Conclusions

This study showed that during the outbreak or initial epidemic phase of COVID-19, from the perspective of the preventive and control personnel, or that was called emergency period, it was possible to control the COVID-19 epidemic within a short period of time by immediately taking active and strict control measures, such as necessary quarantine measures to control the source of infection, cut off the route of transmission, and protect susceptible people, so as to achieve early prevention, early detection, early quarantine, and early treatment. The science and effectiveness of strict control measures during emergency periods were also supported by other research theories and practical evidence.^[41–51]^ At present, relying on factors of temperature and vaccines to control the COVID-19 epidemic will also face many uncertainties, including viral variation,^[52]^ and indecision and oscillation between different vaccination policies may miss the boat and have profound implications for other countries or regions. Different countries or regions may face different levels of epidemics, and their respective processes of passing through the emergency period into the normalized prevention and control period will not be synchronized. It is wise to take active and stringent control measures to intervene in the epidemic according to their own conditions, to quickly pass the emergency period of the epidemic, and to avoid fighting a protracted war during the emergency period.

## Data Availability

All data, models, and code generated or used during the study appear in the submitted article.

## Clinical Trial

NCT12345678

## Authors Declaration

All relevant ethical guidelines have been followed; any necessary IRB and/or ethics committee approvals have been obtained and details of the IRB/oversight body are included in the manuscript.

Yes

All necessary patient/participant consent has been obtained and the appropriate institutional forms have been archived.

Yes

I understand that all clinical trials and any other prospective interventional studies must be registered with an ICMJE-approved registry, such as ClinicalTrials.gov. I confirm that any such study reported in the manuscript has been registered and the trial registration ID is provided (note: if posting a prospective study registered retrospectively, please provide a statement in the trial ID field explaining why the study was not registered in advance).

Yes

I have followed all appropriate research reporting guidelines and uploaded the relevant EQUATOR Network research reporting checklist(s) and other pertinent material as supplementary files, if applicable.

Yes

## Data Availability Statement

All data, models, and code generated or used during the study appear in the submitted article.

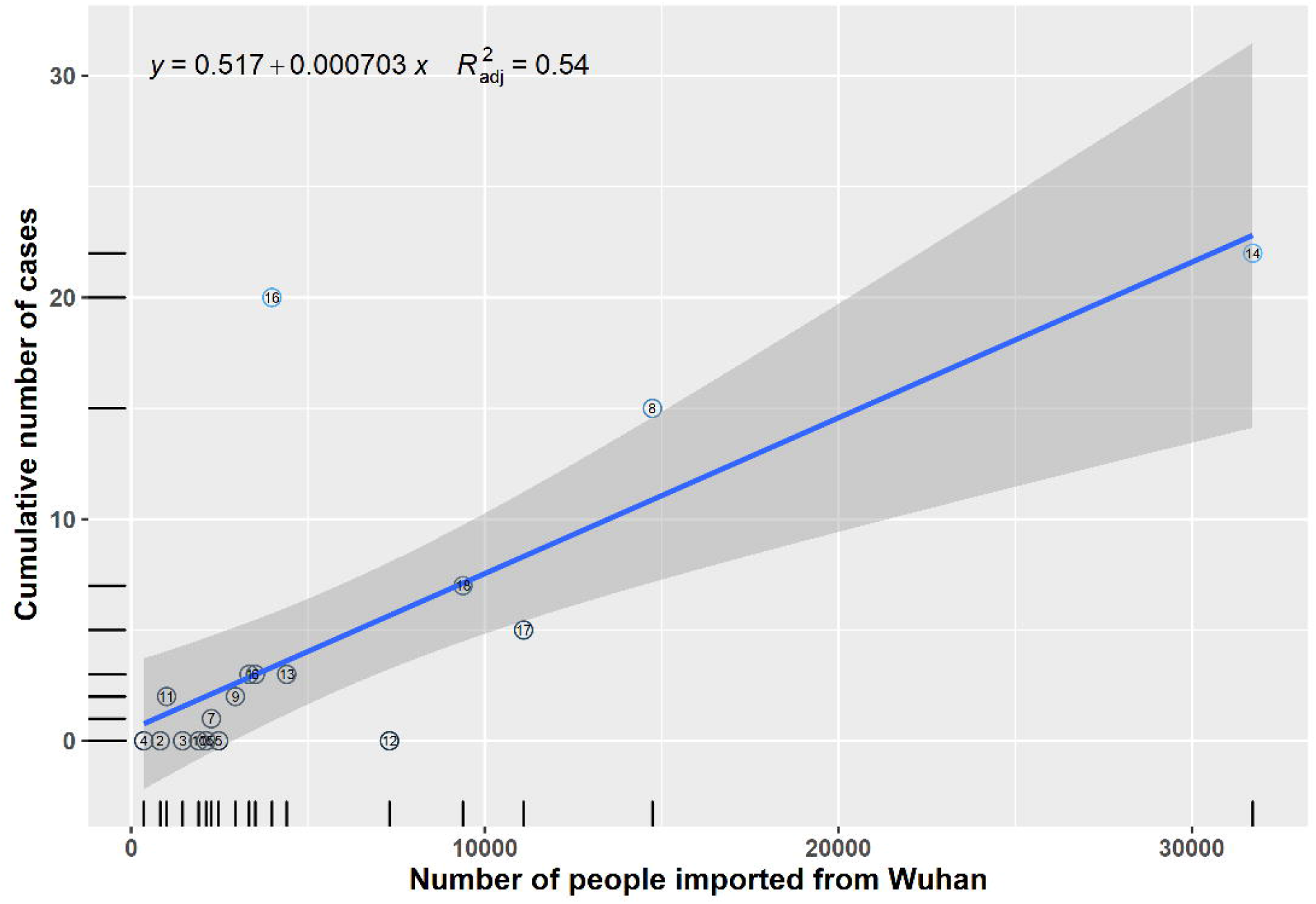

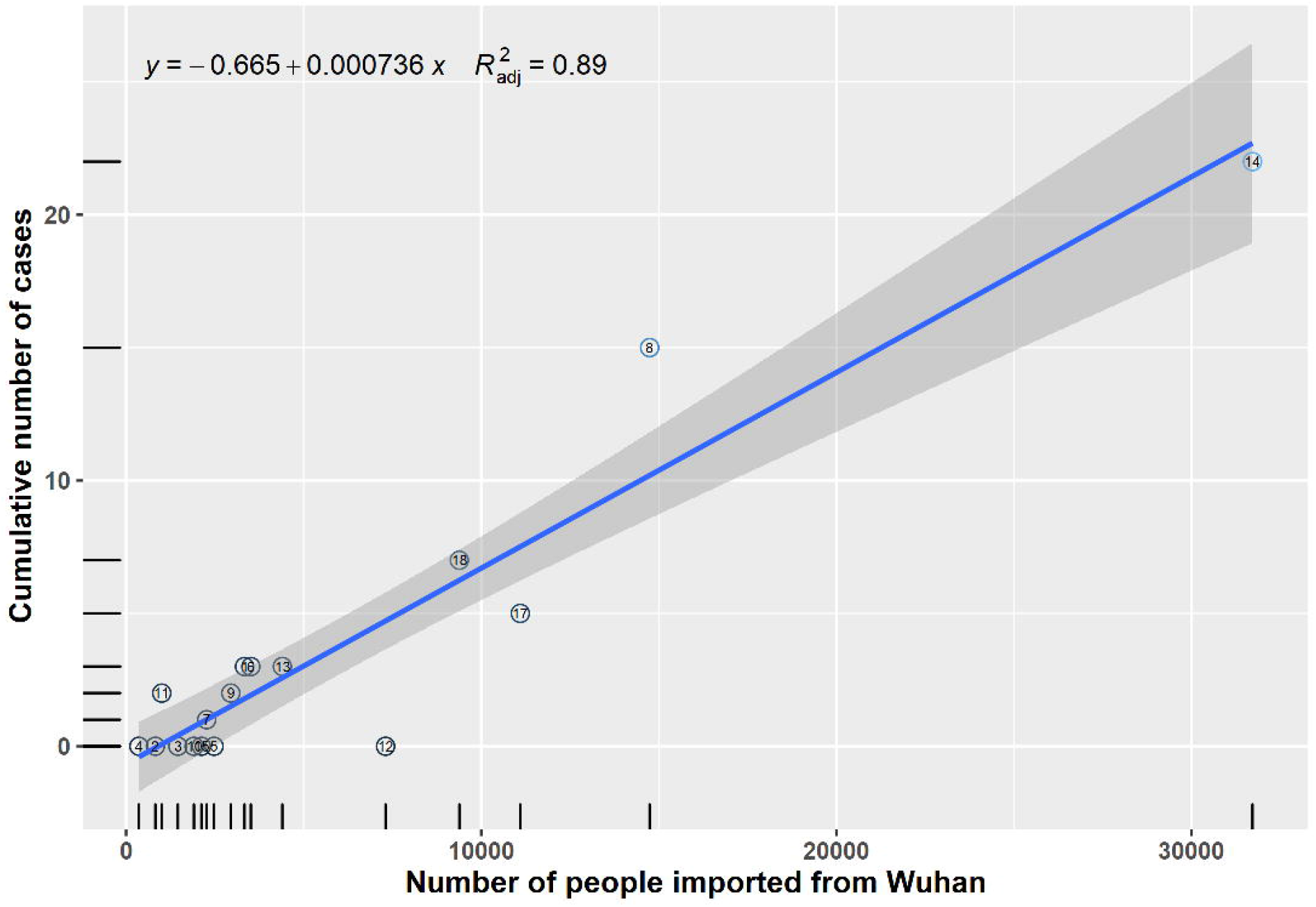

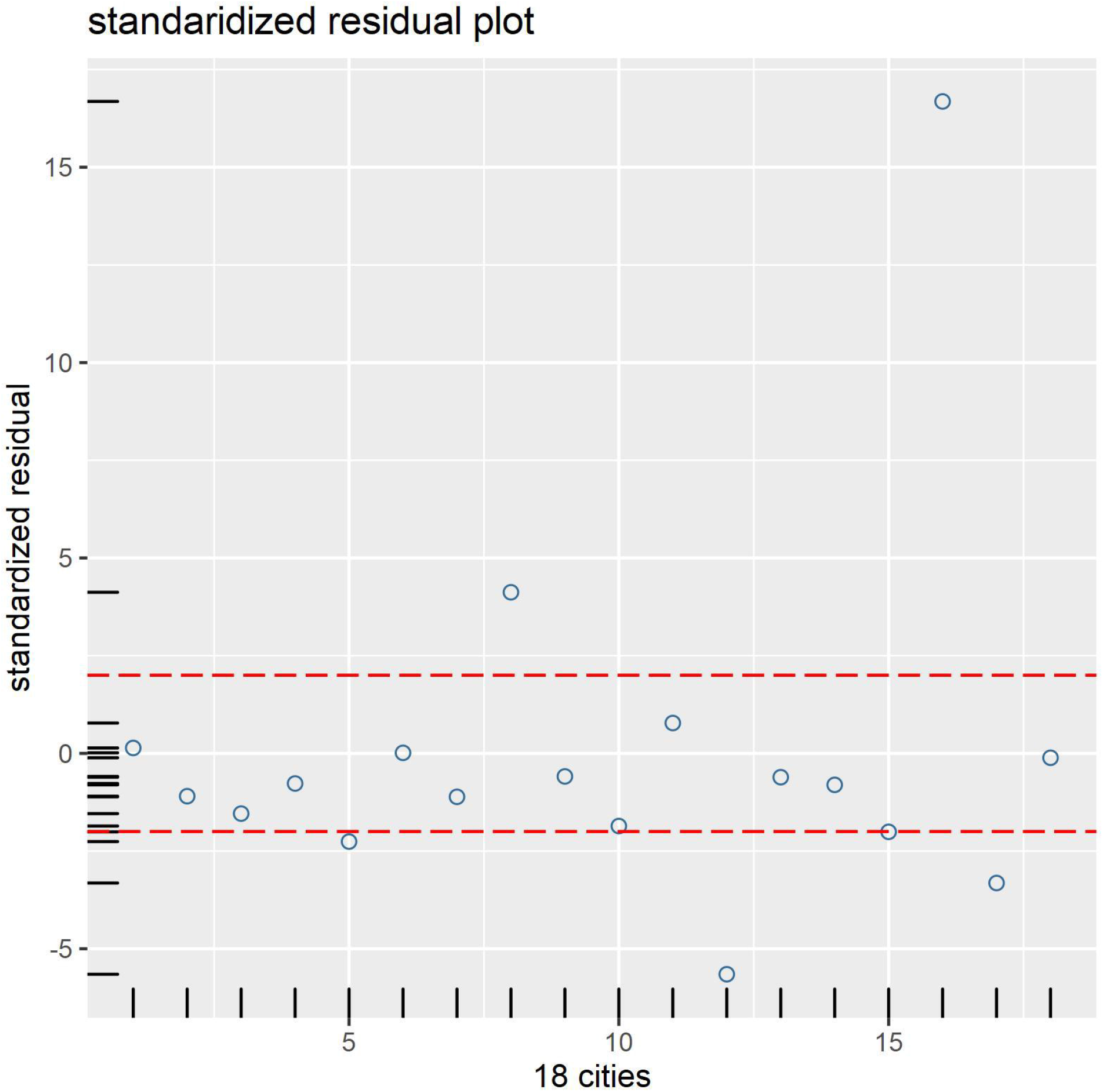

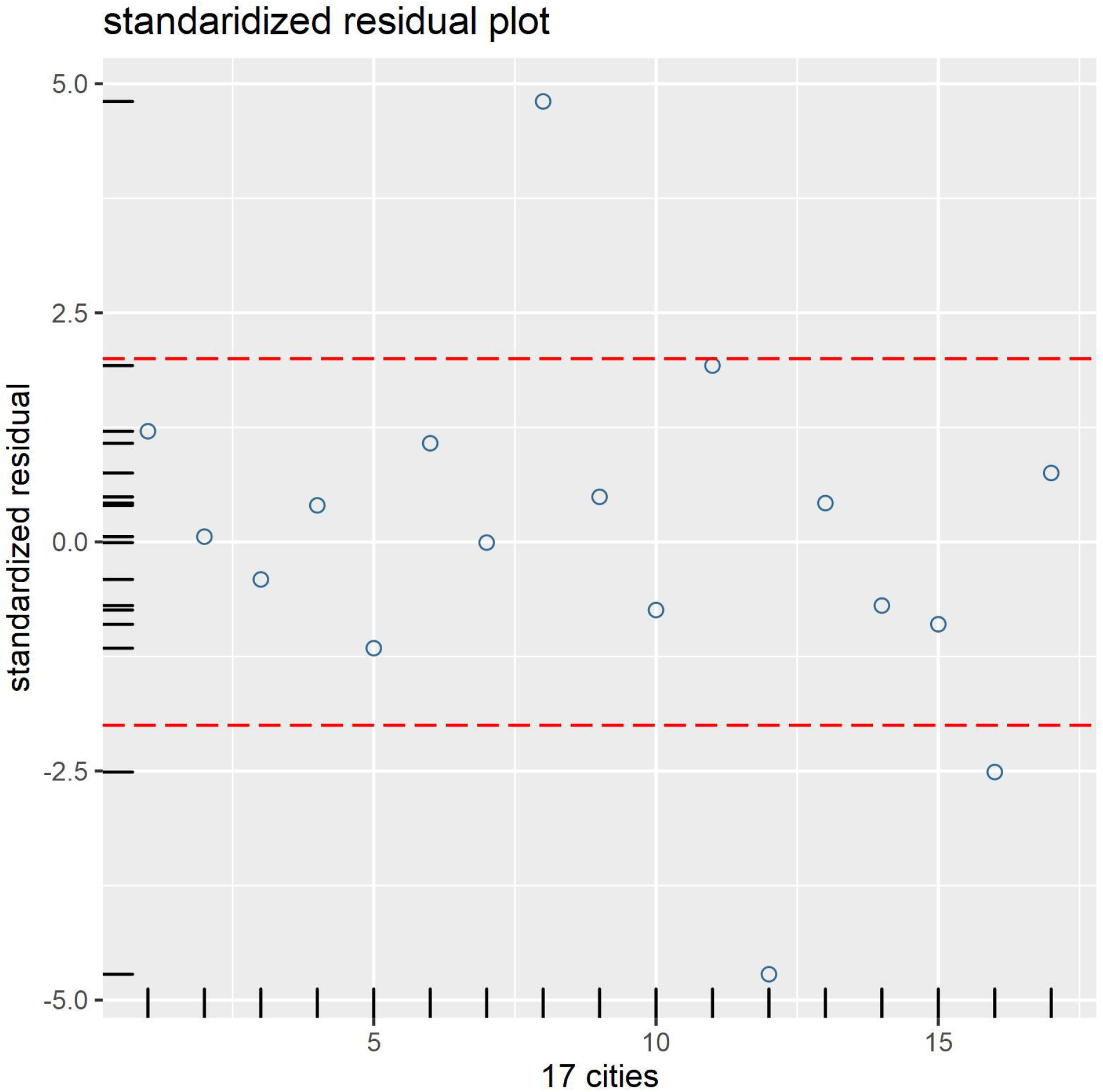

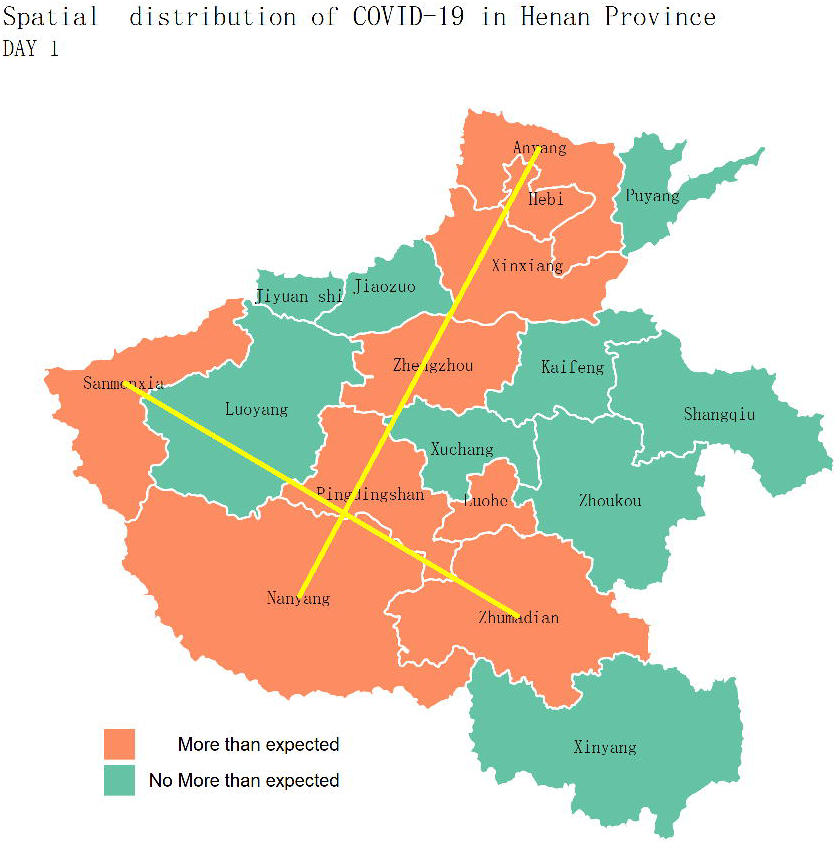

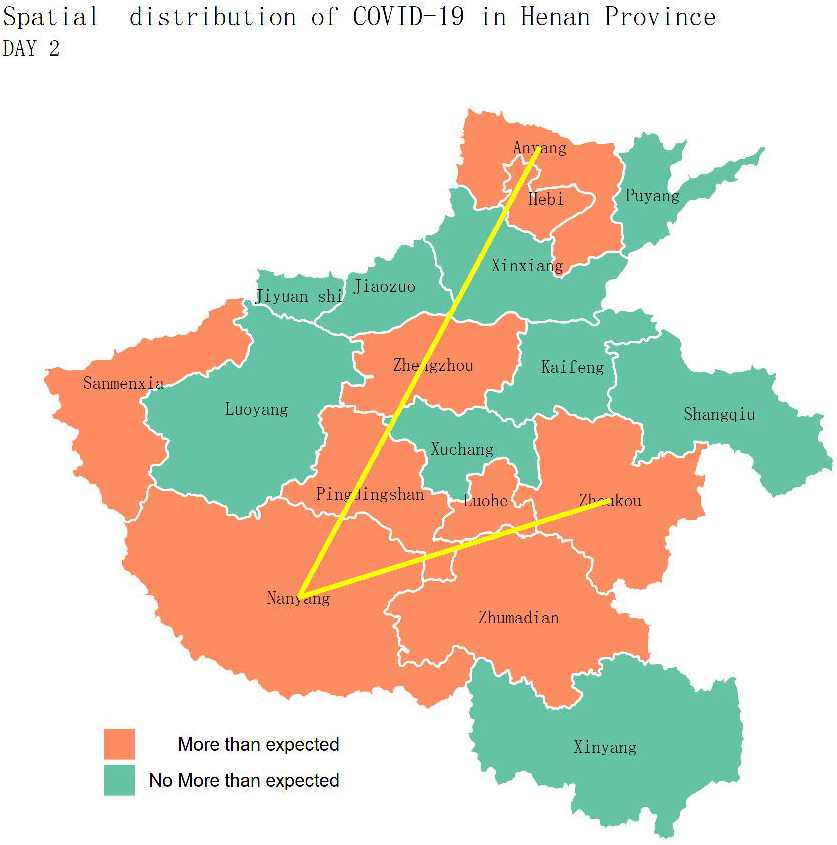

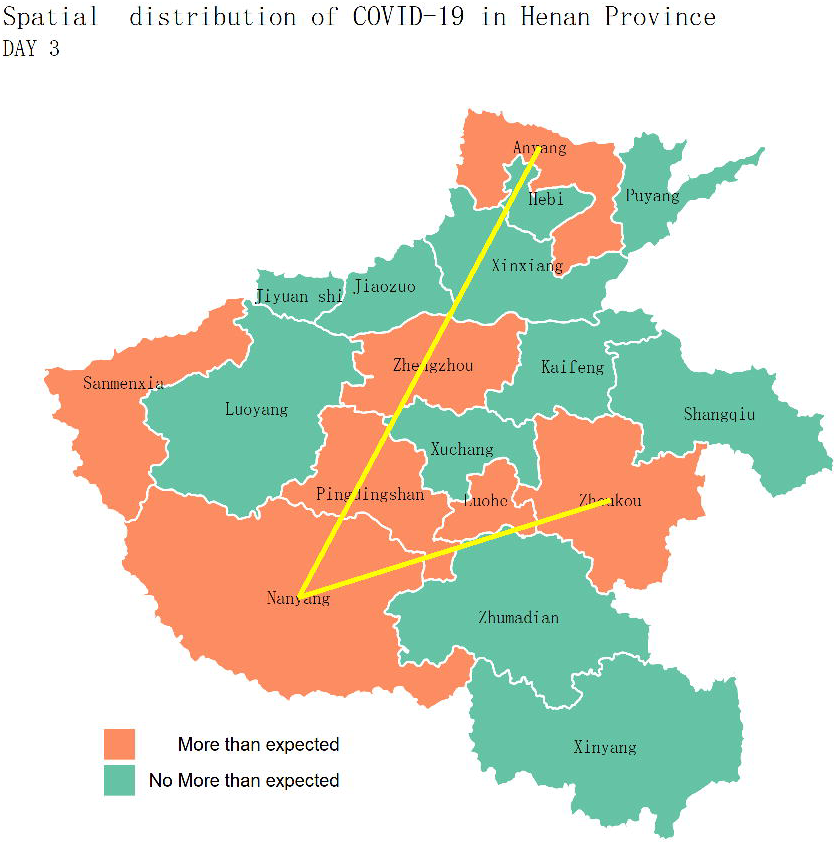

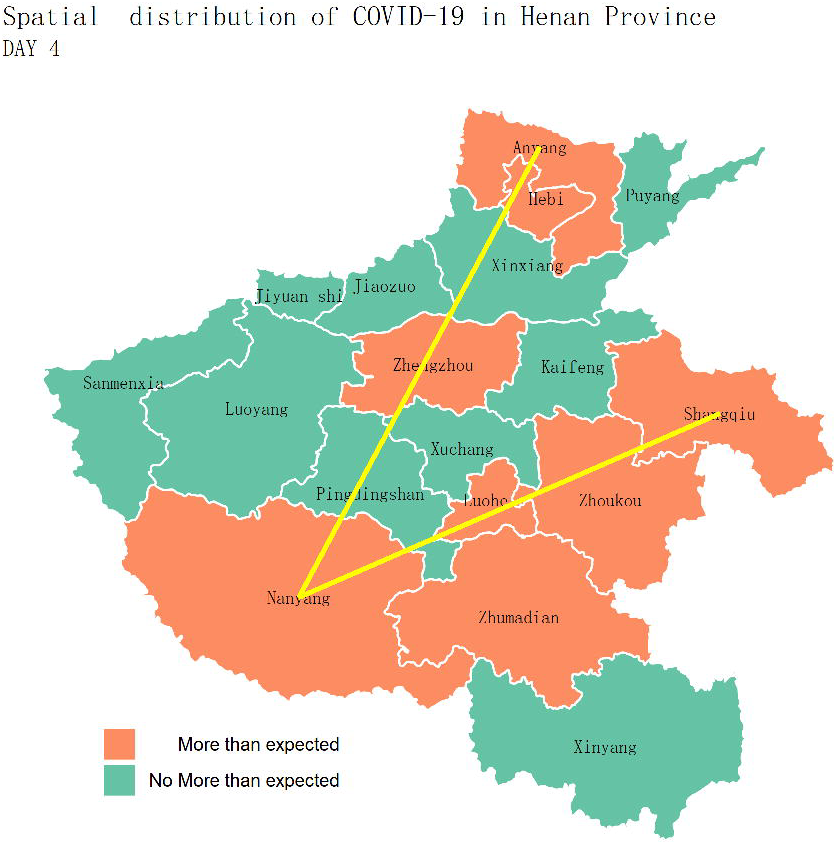

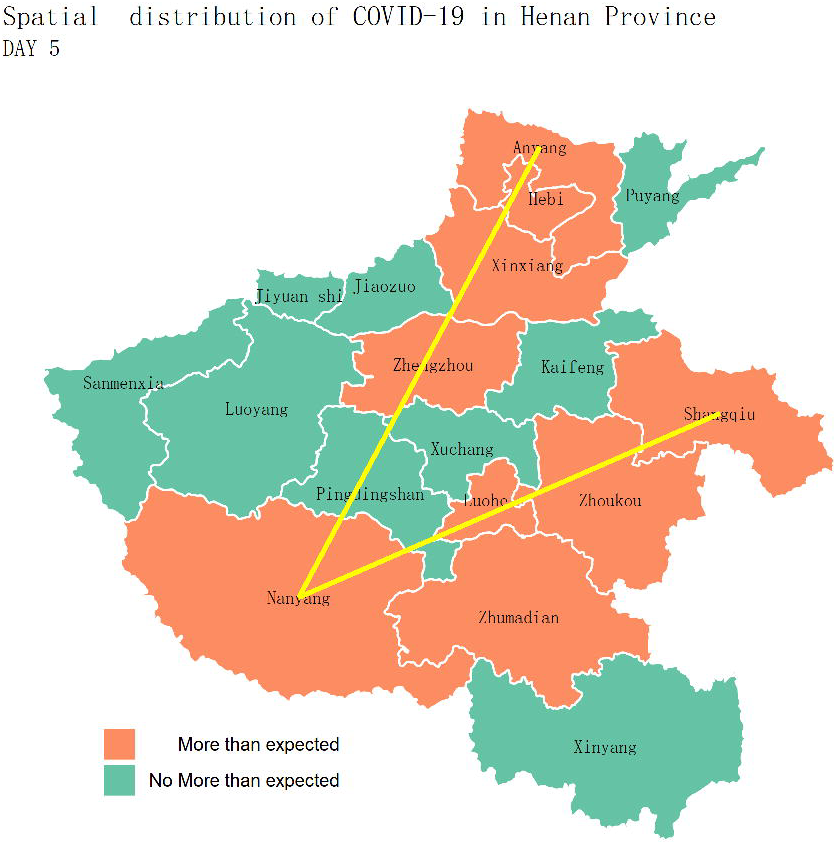

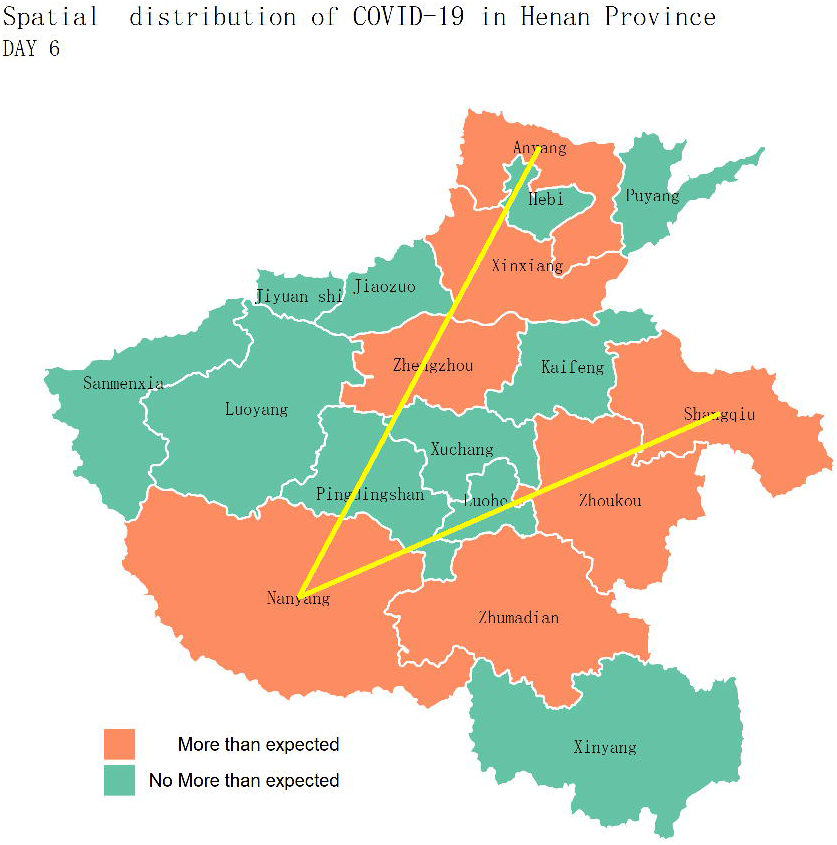

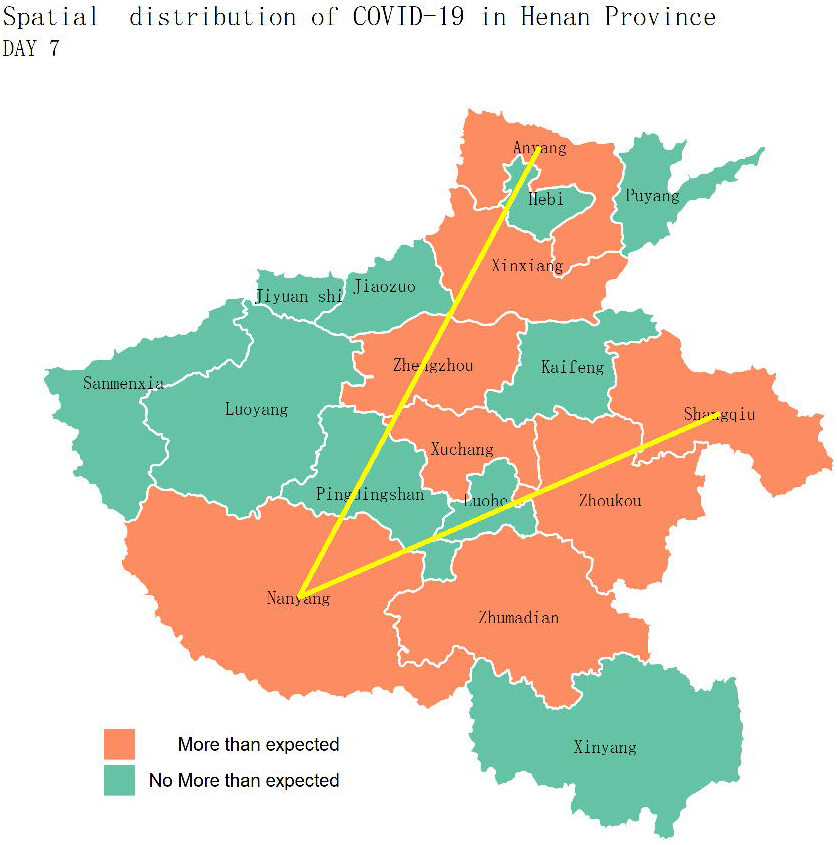

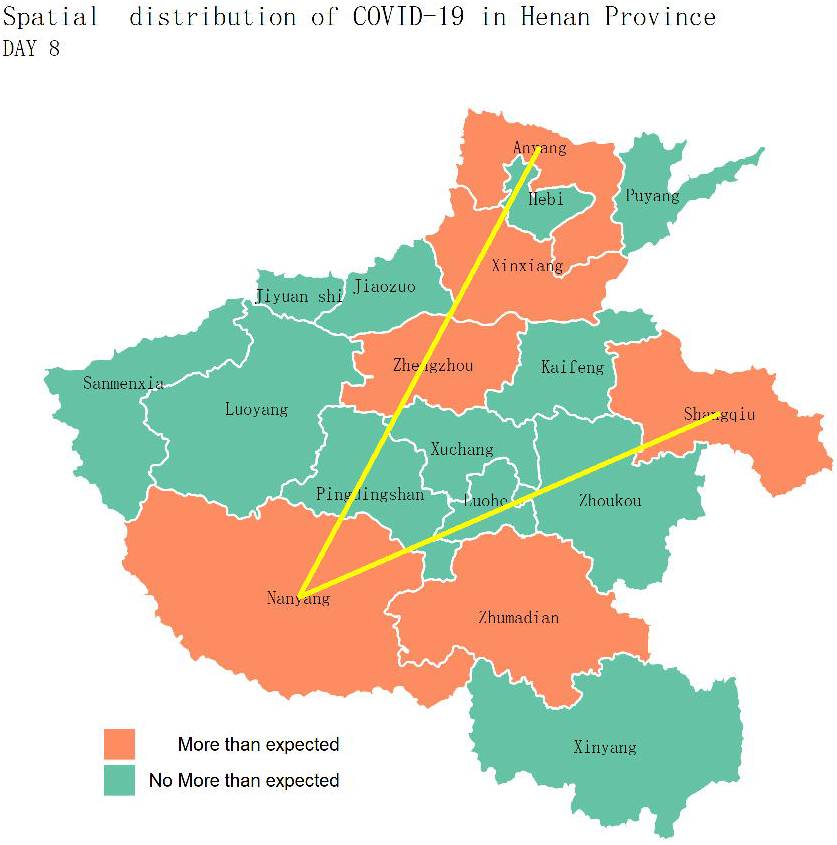

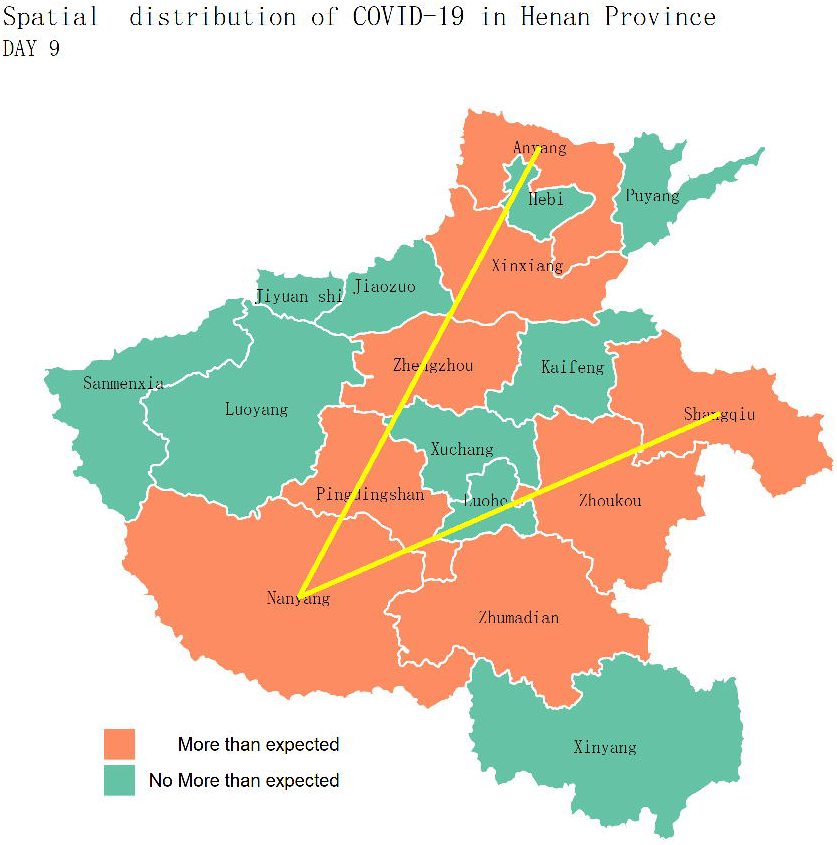

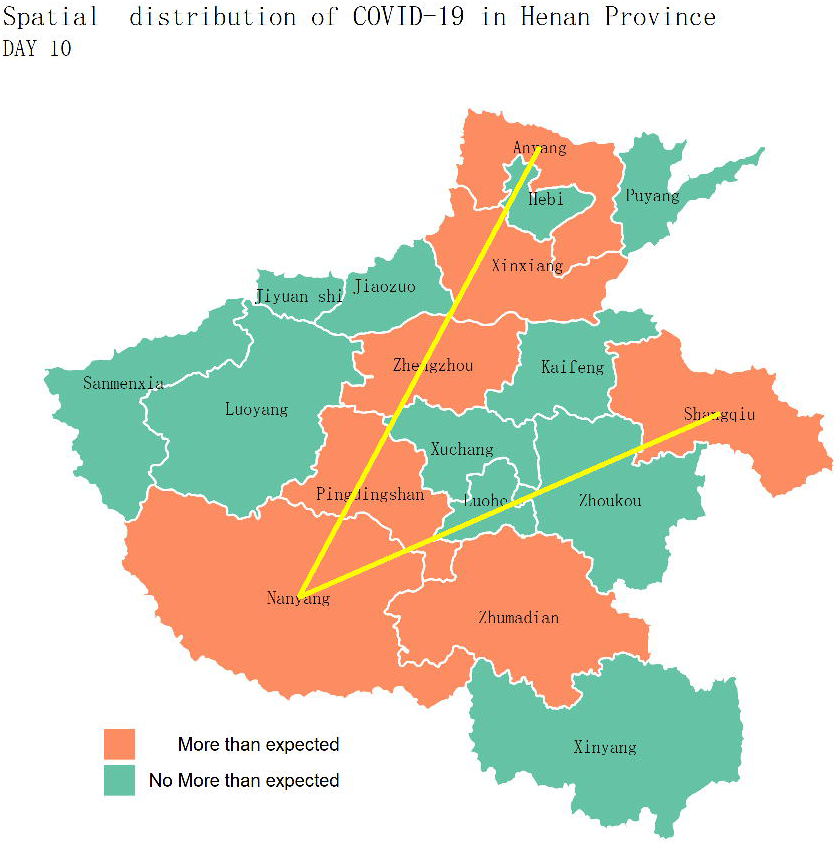

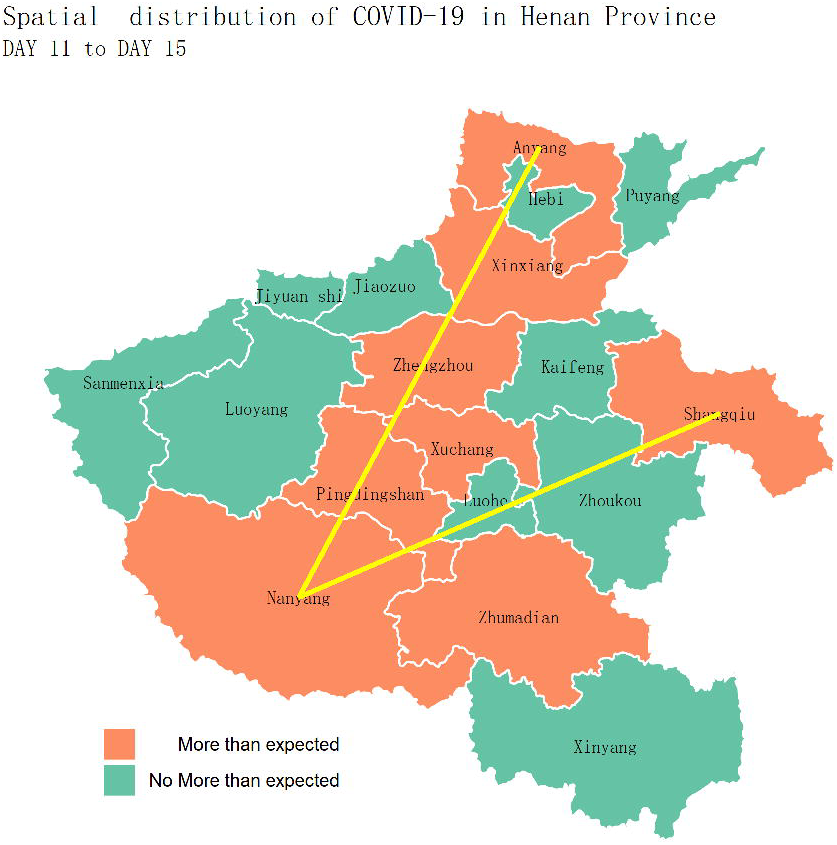

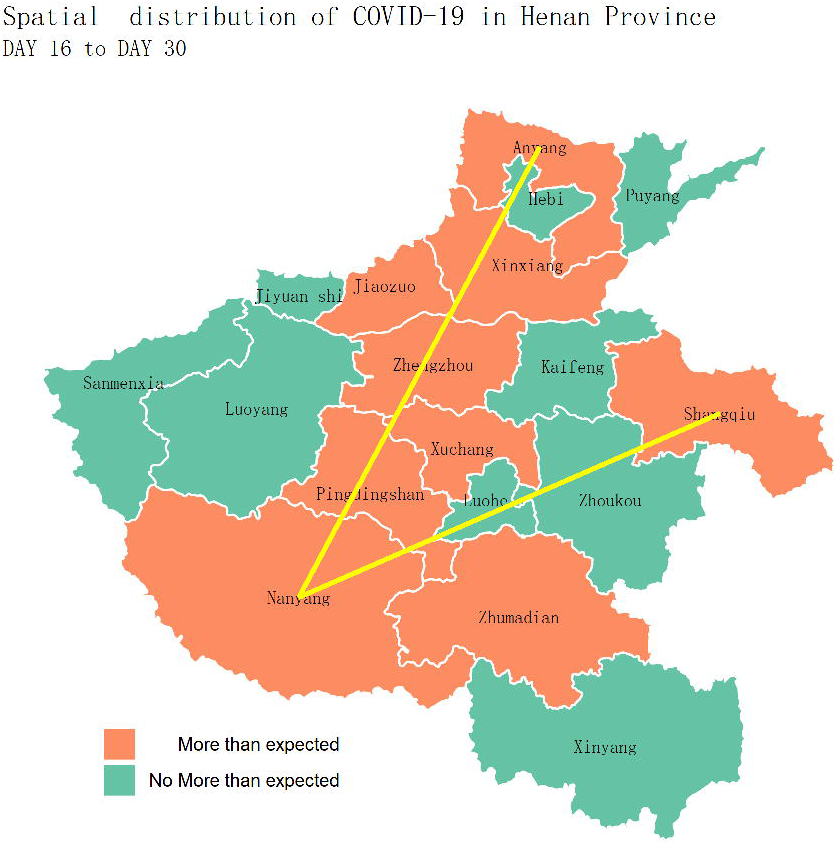

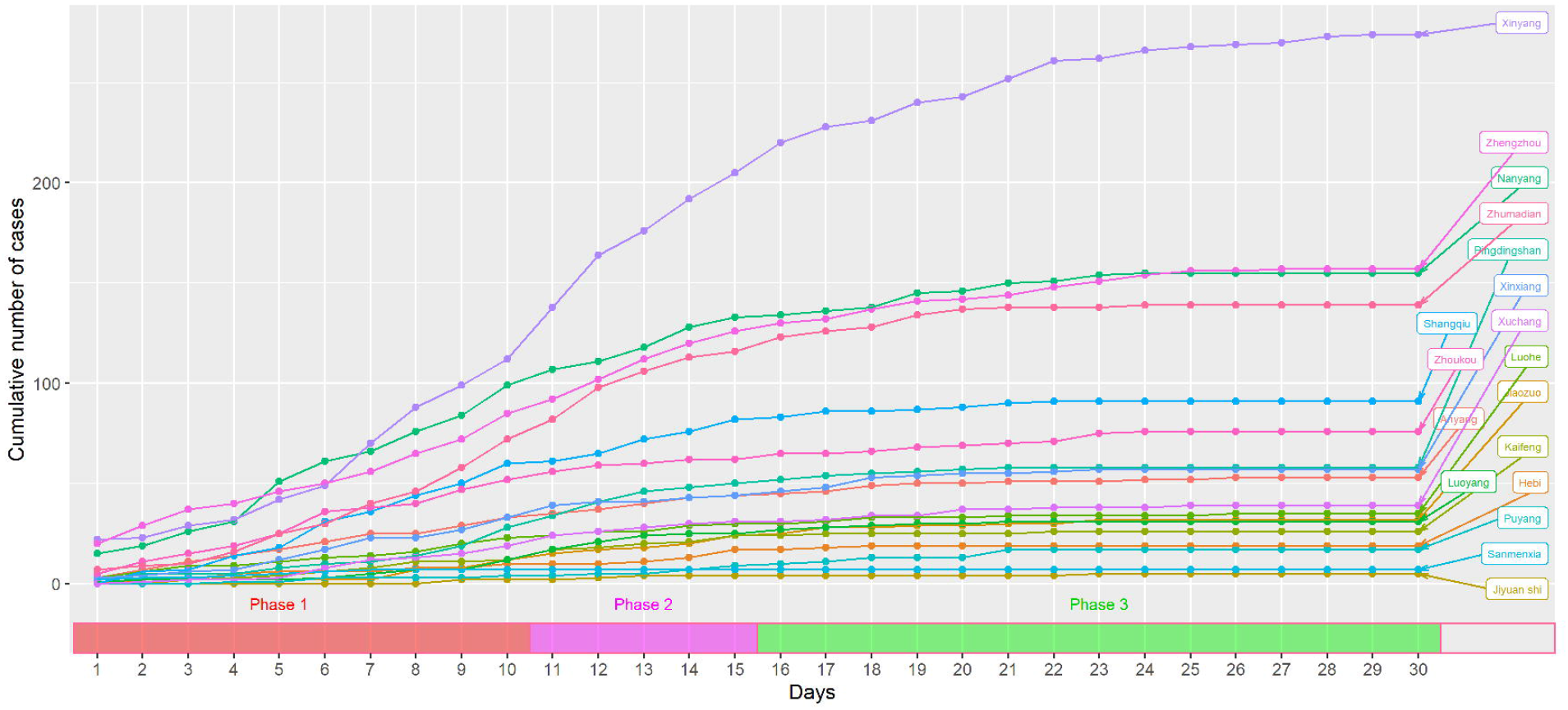

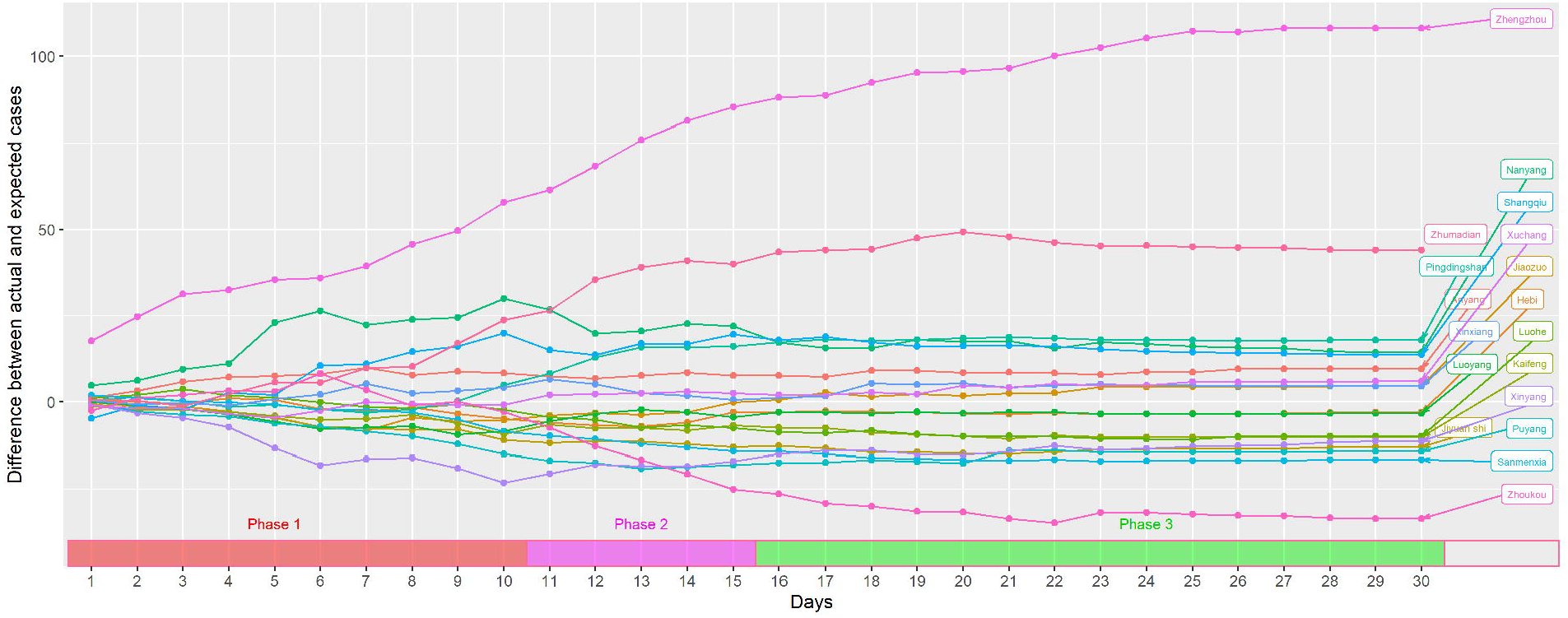

